# Willingness to vaccinate against COVID-19 in the US: Longitudinal evidence from a nationally representative sample of adults from April–October 2020

**DOI:** 10.1101/2020.11.27.20239970

**Authors:** Michael Daly, Eric Robinson

**Affiliations:** Department of Psychology, Maynooth University, Co. Kildare, Ireland; Institute of Population Health Sciences, University of Liverpool, Liverpool, United Kingdom

## Abstract

**Introduction:** Vaccines against COVID-19 have been developed in unprecedented time. However, the effectiveness of any vaccine is dictated by the proportion of the population willing to be vaccinated. In this observational population-based study we examined intentions to be vaccinated against COVID-19 over the course of the pandemic.

**Methods:** We analyzed longitudinal data from a nationally representative sample of 7,547 US adults enrolled in the Understanding America Study (UAS). Participants reporting being willing, undecided and unwilling to get vaccinated against coronavirus across 13 assessments conducted from April-October, 2020. Public attitudes to vaccination against the coronavirus were also assessed.

**Results:** Willingness to vaccinate declined from 71% in April to 53.6% in October. This was explained by an increase in the percentage of participants undecided about vaccinating (from 10.5% to 14.4%) and the portion of the sample unwilling to vaccinate (from 18.5% to 32%). The population subgroups most likely to be undecided/unwilling to vaccinate were those without a degree (undecided: RRR=2.47, 95% CI: 2.04-3.00; unwilling: RRR=1.92, 95% CI: 1.67-2.20), Black participants (undecided: RRR=2.18, 95% CI: 1.73-2.74; unwilling: RRR=1.98, 95% CI: 1.63-2.42), and females (undecided: RRR=1.41, 95% CI: 1.20-1.65; unwilling: RRR=1.29, 95% CI: 1.14-1.46). Those aged 65+, those on high incomes, and other race/ethnicity participants were least likely to be undecided or unwilling to vaccinate. Concerns about potential side effects of a vaccine were common.

**Conclusions:** Intentions to be vaccinated against coronavirus have declined rapidly during the pandemic and close to half of Americans are undecided or unwilling to be vaccinated.

## INTRODUCTION

As of November, 2020 the COVID-19 pandemic has been responsible for more than 1.3 million deaths worldwide^1^. Potential vaccines against COVID-19 have been developed in unprecedented time and early findings suggest there are multiple candidate vaccines that may protect against infection and be suitable for mass roll out in the near future^2,3^. However, the overall effectiveness of any vaccine is dictated, at least in part, by the proportion of the population willing to be vaccinated.. Simulation studies suggest at least three quarters of the population may need to be vaccinated to extinguish the ongoing coronavirus pandemic^4,5^.

During the early stages of the pandemic (March-June), studies of small samples of European and Australian adults suggested that the majority of people surveyed reported that they would be vaccinated when a widely available vaccine was available^6-7^. Similarly, a nationally representative study of adults in China conducted in March found that 9 out of 10 would accept a vaccine when available^8^. US studies conducted early in the pandemic found that between 58% and 86% of adults reported they were likely to be vaccinated against COVID-19^6,9-11^.

However, the rise of ‘fake news’ during the pandemic has been widely acknowledged^11,12^ and widespread misinformation about the pandemic may have been damaging to public uptake of measures designed to reduce the spread of the virus (e.g. mask-wearing, social distancing) and willingness to vaccinate^10,13,14^. In addition, because the speed at which coronavirus vaccines have been developed has been unprecedented and this has been widely reported^2,3^, this may have made the general public more hesitant about accepting a vaccine when available^15,16^. Furthermore, research indicates that in some countries public trust in government handling of the COVID-19 crisis has been negatively affected^17^ and this too may have detrimentally affected intentions to follow public health guidance.

COVID-19 has had a disproportionately large impact on ethnic minorities^18^ and groups from lower socioeconomic backgrounds^19^ and as infections will likely continue to be socially patterned, understanding whether population demographics determine willingness to vaccinate will also be important. For example, research examining previous influenza vaccination programmes has found that vaccination intentions and uptake are reduced among more disadvantaged groups^20,21^. Initial research examining coronavirus vaccination intentions has produced mixed findings on the role of demographic predictors, which may be due to a reliance to date on relatively small and non-representative samples to examine population subgroup differences^9,10,22^.

At present, there is a lack of up-to-date estimates of the proportion of the general population that would be willing to use a vaccine when available and it is unclear whether estimates of vaccination uptake collected much earlier in the pandemic have changed over time. It will be also important to understand whether intentions to vaccinate are socially patterned and more or less likely in specific population sub-groups, in order for public health messages to be directed at those who are least likely to vaccinate^15,16^. In the present research we therefore made use of data from the Understanding America Study (UAS), a large nationally representative panel of US adults who have reported their vaccine intentions on thirteen occasions from the outbreak of the pandemic through to October, 2020.

## METHODS

### Study design and participants

This study utilized data collected as part of the Understanding America Study (UAS), a nationally representative longitudinal study of adults aged 18 and over. The UAS is a probability-based sample recruited via address-based sampling from the US Postal Service Computerized Delivery Sequence file containing all US postal addresses^23^. Participants complete surveys online and those without internet access are provided with tablet computers and internet access. Of 8547 UAS participants eligible to take part in the COVID-19 tracking study, 7547 participated and provided data across 13 waves of assessment conducted every two weeks between April 1^st^ and October 31^st^, 2020^24^.

In total, participants provided 80,060 observations across the 13 survey waves (average response rate of 81.6% among COVID tracking study participants). A small portion (2%) of observations were omitted because they were submitted after October 31^st^ or were missing vaccination intentions or covariate data leaving a total of 78,453 observations (10.4 per participant). The UAS weights were applied to adjust for unequal probabilities of selection into the UAS. Post-stratification weights were also incorporated to provide a correction for non-response by aligning each survey wave with the distribution of demographic characteristics of the US population^25^.

### Measures

In each survey wave participants indicated how likely there were to get vaccinated for coronavirus when a vaccine becomes available to the public on a five-point scale. Participants were classified as either: (1) Undecided (responses of ‘unsure’), (2) Unwilling to vaccinate (responses of somewhat or very *unlikely* to vaccinate), or (3) Willing to vaccinate (responses of somewhat or very *likely* to vaccinate).

Vaccination intentions were predicted by month of survey (April, May, June, July, August, September, October) and a set of demographic variables: age (coded as 18-34, 35-44, 50-64, 65+), sex (coded as male, female), race/ethnicity (White, Hispanic, Black, Other race/ethnicity), household income (≤$40,00/$40,000–$100,000/ ≥$100,000 gross per annum), college degree (vs. none), and the presence of a chronic health condition (present vs. not present). Specifically, participants indicated whether they had been diagnosed with the following conditions: diabetes, cancer, heart disease, kidney disease, asthma, chronic lung disease, an autoimmune disease.

Participants also reported their level of agreement (from 1 = strongly disagree, to 4 = strongly agree) with nine items assessing their attitudes towards a potential vaccine (see Table 3 for items in full) in late October (14^th^-31^st^). Questions assessed participant beliefs that the COVID vaccine would be beneficial, important for personal and community health, and a good way to protect from coronavirus disease. Participants also indicated whether they agreed approved vaccines would be effective and whether they were concerned about the lack of long-term follow-up information and potential side effect of a COVID vaccine were assessed (e.g. “I think the COVID-19 vaccine might cause lasting health problems for me.”).

### Statistical analysis

First, we examined trends in vaccinate intentions over the period of the study by comparing the prevalence of willingness/undecided/unwillingness to vaccinate in April and October, 2020. To estimate the statistical significance of time trends we used multinomial logistic regression analysis with robust standard errors clustered at the individual-level. Those willing to vaccinate were compared to: (i) those undecided on vaccination, and (ii) those unwilling to vaccinate. A series of multinomial logistic regressions were run to identify if the relative risk of being undecided or unwilling to vaccinate increased from April to October for the overall sample and each population subgroup examined. This model contrasts the natural log [Pr(Willing to vaccinate)/Pr(Unwilling to vaccinate)] and natural log [Pr(Undecided on vaccination)/Pr(Unwilling to vaccinate)] estimates across different demographic groups to ascertain relative risk ratios (RRR).

Next, multinomial logistic regression was used to estimate the extent to which survey month and different demographic factors predicted vaccination intentions. A single adjusted analysis was used to estimate the independent effect of each predictor variable (i.e. month of survey, age, sex, race/ethnicity, educational attainment, income, presence of a chronic condition) controlling for all others. In addition, we tested a separate model where interactions between survey month and participant demographics were added to our main model to test whether changes in vaccination intentions over time differed systematically between demographic groups.

All analyses incorporated the UAS sampling weights to generate nationally representative estimates. RRRs and 95% CIs were estimated using the Stata version 15 (Statacorp).

## RESULTS

Participants were aged 47.2 (SD = 16.6) years on average, 52.1% were female, 34.2% had a college degree, and 64.1% were White, 17.8% Hispanic, 12.2% Black, and 5.9% Other race/ethnicity (see Table 1). On average, willingness to vaccinate declined from 71% in April to 53.6% in October. This was explained by an increase in the percentage of participants undecided about vaccinating against COVID-19 (from 10.5% to 14.4%) and the portion of the sample unwilling to vaccinate (from 18.5% to 32%), as shown in Table 1. A decrease in the willingness to vaccinate against COVID-19 between April and October was evident across all population subgroups examined (Table 1).

**Table 1.** Sample characteristics of participants in the Understanding American Study (UAS; N = 7,547, Obs. = 78,453) and vaccination intentions in April and October, 2020.

|  |  | Willing to<br>vaccinate<sup>a</sup> |  | Undecided on<br>vaccination<sup>a</sup> |  | Unwilling to<br>vaccinate<sup>a</sup> |  |
| --- | --- | --- | --- | --- | --- | --- | --- |
| Survey month |  | April | October | April | October | April | October |
| Variable | Sample size (%) | % | % | % | % | % | % |
| Overall sample |  | 71.0 | 53.6 | 10.5 | 14.4 | 18.5 | 32.0 |
| Age group |  |  |  |  |  |  |  |
| 18-34 | 2024 (26.8) | 65.6 | 47.2 | 12.9 | 16.2 | 21.5 | 36.6 |
| 35-49 | 2305 (30.5) | 67.5 | 49.6 | 11.5 | 15.5 | 21.0 | 34.9 |
| 50-64 | 1832 (24.3) | 73.1 | 54.2 | 10.6 | 15.1 | 16.3 | 30.7 |
| 65+ | 1387 (18.4) | 79.9 | 65.9 | 6.0 | 10.0 | 14.1 | 24.0 |
| Sex |  |  |  |  |  |  |  |
| Male | 3613 (47.9) | 75.1 | 60.0 | 8.2 | 11.7 | 16.7 | 28.3 |
| Female | 3934 (52.1) | 67.2 | 47.6 | 12.7 | 17.0 | 20.1 | 35.4 |
| Race/ethnicity |  |  |  |  |  |  |  |
| White | 4840 (64.1) | 74.7 | 57.3 | 8.6 | 13.0 | 16.7 | 29.7 |
| Hispanic | 1345 (17.8) | 67.4 | 47.5 | 12.2 | 16.2 | 20.5 | 36.3 |
| Black | 917 (12.2) | 47.9 | 33.8 | 22.0 | 21.8 | 30.1 | 44.3 |
| Other race/<br>ethnicity | 445 (5.9) | 86.5 | 68.6 | 4.0 | 11.1 | 9.5 | 20.4 |
| Education |  |  |  |  |  |  |  |
| No degree | 4970 (65.8) | 65.7 | 45.1 | 13.1 | 18.2 | 21.2 | 36.7 |
| College degree | 2578 (34.2) | 81.4 | 68.9 | 5.4 | 7.8 | 13.2 | 23.4 |
| Income level<sup>b</sup> |  |  |  |  |  |  |  |
| Low income | 2884 (38.2) | 64.1 | 43.6 | 15.6 | 21.4 | 20.3 | 35.0 |
| Middle income | 3007 (40.4) | 71.6 | 55.0 | 8.6 | 12.1 | 19.8 | 33.0 |
| High income<sup>b</sup> | 1656 (21.9) | 81.1 | 66.9 | 5.6 | 7.8 | 13.3 | 25.4 |
| Chronic condition<sup>c</sup> |  |  |  |  |  |  |  |
| No | 5060 (67.0) | 69.5 | 52.5 | 10.6 | 14.2 | 19.9 | 33.4 |
| Yes | 2446 (32.4) | 74.1 | 55.8 | 10.3 | 14.9 | 15.6 | 29.2 |

Note: Weighted demographic characteristics and vaccination intentions are presented.
<sup>a</sup> Based-on responses to the question “How likely are you to get vaccinated for coronavirus once a vaccine is available to the public?”
<sup>b</sup> Households earning less than \$40,000 a year classified as low income, those earning \$40,000 - \$100,000 middle income, and those above this threshold as high-income.
<sup>c</sup> Diagnosed with any of the following: diabetes, cancer, heart disease, kidney disease, asthma, chronic lung disease, an autoimmune disease. 41 participants did not provide chronic illness data.

An unadjusted multinomial logistic regression analysis confirmed that from April to October, 2020 there was a statistically significant higher risk of being undecided (RRR = 1.82, 95% CI: 1.62-2.05) or unwilling (RRR = 2.29, 95% CI: 2.11-2.48) to be vaccinated versus being willing to get vaccinated (see Table S1). Unadjusted multinomial logistic regression analyses also showed that all population subgroups were more likely to be undecided or unwilling to vaccinate in October compared to April (Table S1). There was also an over 2-fold higher relative likelihood of being undecided or unwilling to get the COVID-19 vaccine in October compared to April, 2020 (undecided: RRR = 2.03, 95% CI: 1.79-2.29; unwilling: RRR = 2.47, 95% CI: 2.27-2.68) in a fully adjusted model that included controls for participant demographic factors and the presence of chronic illness (Table 2). An examination of month-to-month changes confirmed that the likelihood of being undecided or unwilling to vaccinate (versus being willing to vaccinate) increased in a graded fashion from April to October, as shown in Table 2 and illustrated in Figure 1.

**Table 2.** Results of adjusted multinomial logistic regression analyses examining demographic predictors and temporal changes in indecision and unwillingness to vaccinate against COVID-19 in the United States (N = 7,547, Obs. = 78,453).

| Variable | Undecided on vaccination |  | Unwilling to vaccinate |  |
| --- | --- | --- | --- | --- |
|  | RRR<sup>a</sup> | (95% CI) | RRR<sup>a</sup> | (95% CI) |
| Month (ref. is April) |  |  |  |  |
| May | 1.27*** | (1.15, 1.41) | 1.54*** | (1.44, 1.66) |
| June | 1.34*** | (1.18, 1.52) | 1.65*** | (1.51, 1.80) |
| July | 1.47*** | (1.30, 1.67) | 1.76*** | (1.62, 1.91) |
| August | 1.50*** | (1.33, 1.68) | 1.97*** | (1.82, 2.13) |
| September | 1.92*** | (1.70, 2.16) | 2.34*** | (2.16, 2.55) |
| October | 2.03*** | (1.79, 2.29) | 2.47*** | (2.27, 2.68) |
| Age group (ref. is 18-34) |  |  |  |  |
| 35-49 | 1.07 | (0.87, 1.33) | 1.02 | (0.85, 1.21) |
| 50-64 | 0.88 | (0.71, 1.09) | 0.81* | (0.68, 0.97) |
| 65+ | 0.49*** | (0.38, 0.63) | 0.61*** | (0.51, 0.74) |
| Sex (ref. is male) |  |  |  |  |
| Female | 1.41*** | (1.20, 1.65) | 1.29*** | (1.14, 1.46) |
| Race/ethnicity (ref. is White) |  |  |  |  |
| Hispanic | 1.05 | (0.82, 1.35) | 1.02 | (0.84, 1.25) |
| Black | 2.18*** | (1.73, 2.74) | 1.98*** | (1.63, 2.42) |
| Other race/ethnicity | 0.57** | (0.40, 0.82) | 0.52*** | (0.39, 0.71) |
| Education (ref. is degree) |  |  |  |  |
| No degree | 2.47*** | (2.04, 3.00) | 1.92*** | (1.67, 2.20) |
| Income level (ref. is low income)<sup>b</sup> |  |  |  |  |
| Middle income | 0.58*** | (0.48, 0.69) | 1.01 | (0.88, 1.16) |
| High income | 0.40*** | (0.32, 0.50) | 0.75** | (0.63, 0.90) |
| Chronic condition<sup>c</sup> | 0.96 | (0.81, 1.14) | 0.84* | (0.74, 0.96) |

<sup>a</sup> Estimates are relative risk ratios derived from multinomial logistic regression with standard errors adjusted for clustering at the individual-level and controlling for all characteristics presented. For all analyses “willing to vaccinate” was the outcome reference group.
<sup>b</sup> Households earning less than \$40,000 a year classified as low income, those earning \$40,000 - \$100,000 middle income, and those above this threshold as high-income.
<sup>c</sup> Diagnosed with any of the following: diabetes, cancer, heart disease, kidney disease, asthma, chronic lung disease, an autoimmune disease.
\* $P < .05$ . \*\* $P < .01$ . \*\*\* $P < .001$

**Figure 1.**
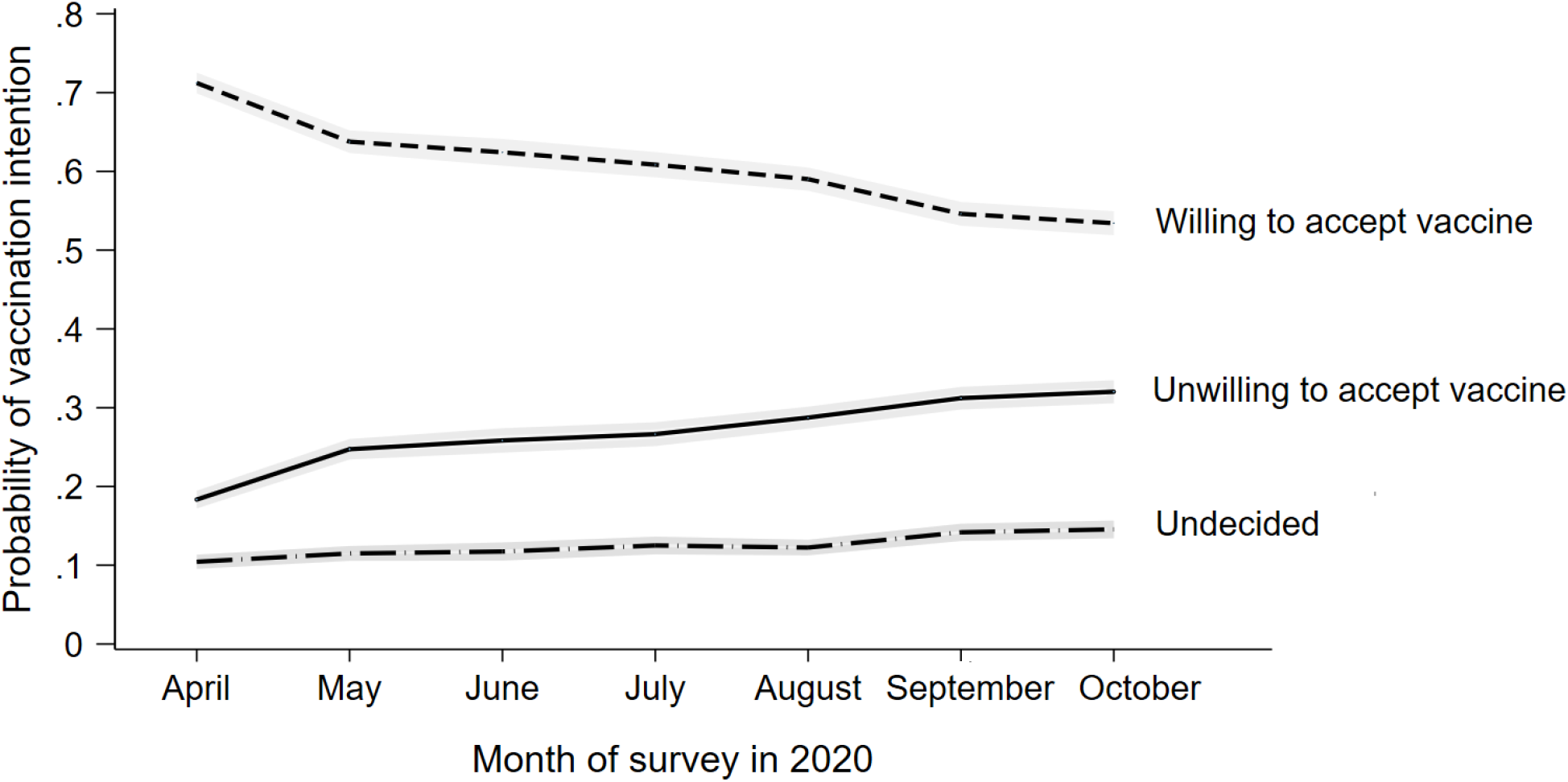
Change in vaccination intentions across 13 waves of the Understanding America Study conducted between April 1^st^ and October 31^st^, 2020. Graph is based on an analysis of 78453 observations on 7547 participants. Estimates are predicted probabilities from marginal effects calculated after a multinomial logistic regression model adjusted for age, sex, race/ethnicity, household income, educational attainment, and the presence of pre-existing health conditions. 95% confidence intervals are presented in grey.

When all observations from 13 survey waves were examined, those without a college degree were at elevated relative risk of being undecided or unwilling to vaccinate (undecided: RRR = 2.47, 95% CI: 2.04-3.00; unwilling: RRR = 1.92, 95% CI: 1.67-2.20), as were Black participants (undecided: RRR = 2.18, 95% CI: 1.73-2.74; unwilling: RRR = 1.98, 95% CI: 1.63-2.42) and females (undecided: RRR = 1.41, 95% CI: 1.20-1.65; unwilling: RRR = 1.29, 95% CI: 1.14-1.46). In contrast, a reduced relative risk of being undecided or unwilling to vaccinate was found among those aged 65+ (undecided: RRR = 0.49, 95% CI: 0.38-0.63 unwilling: RRR=0.61, 95% CI: 0.51-0.74), those on high household incomes (undecided: RRR = 0.40, 95% CI: 0.32-0.50; unwilling: RRR=0.52, 95% CI: 0.39-0.71), and other race/ethnicity participants (undecided: RRR = 0.57, 95% CI: 0.40-0.82; unwilling: RRR=0.52, 95% CI: 0.39-0.71).

An examination of the interactions between survey month and individual demographic characteristics did not yield evidence for systematic differences in changes in vaccination intentions over time between demographic groups.

Finally, we examined attitudes towards the vaccine reported between October 14-31, 2020. The majority of the sample (70-80%) agreed that the COVID vaccine would be personally beneficial, important for personal and community health, a good way to protect from coronavirus disease, and effective if approved by the U.S. Food and Drug Administration (FDA) or the Centre for Disease Control and Prevention (CDC; see Table 3). However, responses differed markedly between those willing and unwilling to be vaccinated. For example, while 92% of those who were willing to be vaccinated agreed that the vaccine would be effective if approved by the FDA or CDC, only 43% of those unwilling to be vaccinated agreed. In the overall sample it was common for participants to report concerns over the vaccine and 69.7% agreed they were concerned about serious side effects of the vaccine. Forty-four % agreed the vaccine might cause lasting health problems for them. However, such concerns were more prevalent among those unwilling to be vaccinated. For example, 65% of this group were concerned about lasting health problems resulting from the vaccine compared to 27% of those willing to be vaccinated.

**Table 3.** Attitudes Towards Vaccination against COVID-19 in the Understanding America Study assessed between October 14^th^ and 31^st^ 2020 (N = 5762).

| Question<sup>b</sup> | Full sample |  | Willing to vaccinate<sup>a</sup> |  | Undecided on vaccination<sup>a</sup> |  | Unwilling to vaccinate<sup>a</sup> |  |
| --- | --- | --- | --- | --- | --- | --- | --- | --- |
|  | Agree (%) | Disagree (%) | Agree (%) | Disagree (%) | Agree (%) | Disagree (%) | Agree (%) | Disagree (%) |
| 1. The COVID vaccine will be important for my health. | 71.2 | 28.8 | 93.8 | 6.2 | 69.1 | 30.9 | 33.7 | 66.3 |
| 2. Getting a COVID vaccine would be a good way to protect me from coronavirus disease. | 74.2 | 25.8 | 95.4 | 4.6 | 71.6 | 28.4 | 39.3 | 60.7 |
| 3. The COVID vaccine will be effective if it is approved by the FDA or CDC. | 73.7 | 26.3 | 92.4 | 7.6 | 71.0 | 29.0 | 43.1 | 56.9 |
| 4. Getting the COVID vaccine will be important for the health of others in my community. | 79.4 | 20.6 | 96.1 | 3.9 | 77.7 | 22.3 | 51.7 | 48.3 |
| 5. The COVID vaccine will be beneficial to me. | 73.8 | 26.2 | 95.1 | 4.9 | 74.3 | 25.7 | 37.4 | 62.6 |
| 6. I will do what my doctor or health care provider recommends about the COVID vaccine. | 74.5 | 25.5 | 92.8 | 7.2 | 70.9 | 29.1 | 44.7 | 55.3 |
| 7. The COVID vaccine will not be around long enough to be sure it is safe. | 48.1 | 51.9 | 39.9 | 60.1 | 58.8 | 41.2 | 57.3 | 42.7 |
| 8. I am concerned about serious side effects of the COVID vaccine. | 69.7 | 30.3 | 60.6 | 39.4 | 81.4 | 18.6 | 80.2 | 19.8 |
| 9. I think the COVID vaccine might cause lasting health problems for me. | 43.6 | 56.4 | 26.6 | 73.4 | 61.5 | 38.5 | 65.0 | 35.0 |

Note: Estimates are based on weighted data. <sup>a</sup>Based-on responses to the question “*How likely are you to get vaccinated for coronavirus once a vaccine is available to the public?*”. In this survey wave (responses between October 14-31, 2020) 54% of the sample were classified as ‘Willing to vaccinate’, 14% ‘Undecided’, and 32% ‘Unwilling to vaccinate’. <sup>b</sup> Each item was rated on a four-point scale with those responding ‘Somewhat’ or ‘Strongly’ agree are coded as ‘Agree’ and those responding ‘Somewhat’ or ‘Strongly’ disagree are coded as ‘Disagree’.

## DISCUSSION

In a large nationally representative sample of US adults, intentions to be vaccinated against COVID-19 have declined from a high of 71% of the population in April to close to only 54% reporting being willing to vaccinate in October, 2020. Reporting being undecided or unwilling to vaccinate was more likely among those with lower levels of education and income, females, Black (African American) and younger adults. Concerns about the vaccine causing long lasting health problems and uncertainty about the benefits of the vaccine were also common.

Based on estimates that vaccination coverage of close to 75% may be required to vaccinate to eradicate coronavirus^4,5,10,26^, our estimates that close to 50% of the population may be willing to vaccinate are concerning. It will now be critical to better understand the reasons why a large proportion of the population are sceptical about vaccination against COVID-19. Public concerns about the safety of vaccines may be an important driver of the increase in the proportion of the population reporting being unsure or explicitly stating they will not vaccinate^10^.

In line with this, 70% of the present sample reported being concerned about serious side effects of the vaccine and 44% believed that a vaccine might cause lasting health problems for them. To some extent these concerns are to be expected given the unprecedented speed at which vaccines have been developed^2,3^ and current lack of information on long-term safety of candidate vaccines. However, the rise of anti-vaccination misinformation (e.g. misleading healthcare information, conspiracy theories) may also have played a role in explaining this increase^11-14,26^. It will therefore be critical that accurate safety information is widely and transparently communicated by trusted sources to promote confidence in the scientific decision-making underpinning the approval of COVID-19 vaccines^16,21^.

It will also be important to address social inequalities in vaccination intentions^20,21^, and to ensure widespread uptake of effective COVID-19 vaccines. In the present study, willingness to vaccinate was strikingly lower among more disadvantaged groups (e.g. African Americans, those with lower income and education levels) and these groups have already been disproportionately affected by COVID-19^18,19^. Previous research on influenza vaccines also suggests that access issues may prevent minority and disadvantaged groups from being vaccinated^21,27^. It will therefore be important to identify strategies to reduce social inequalities in both vaccination intentions and opportunities to vaccinate^20^.

Strengths of the present research include the use of a large probability-based nationally representative sample of adults allowing generalizations to be made to the population. The current study also moves beyond prior research by including a high frequency longitudinal assessment of vaccination intentions throughout the pandemic. In addition, participant concerns and perceptions of the benefits of a potential vaccine were assessed to shed light on potential reasons for low willingness to vaccinate.

Limitations are our reliance on self-reported intentions and the lack of detailed assessment of factors that may explain why vaccination intentions have declined over time in the US. However, in advance of the deployment of a COVID vaccine it is necessary to rely on intention-based measures which have been shown to predict vaccination behavior^27^. Further, intentions are malleable and represent a target for evidence-based approaches aiming to increase the proportion of the population that are willing to vaccinate^28^. This now represents an urgent public health priority to minimize further loss of life due to the COVID-19 pandemic^29^. Finally, studies tracking vaccination attitudes including perceived health benefits and side-effects of vaccination, long-term health concerns, and the role of misinformation are now needed to provide an in-depth understanding of the drivers of changes in vaccine intentions.

## CONCLUSION

Intentions to be vaccinated against coronavirus have declined rapidly during the pandemic and close to half of Americans are undecided or unwilling to be vaccinated. This reduced willingness to vaccinate may undermine the pandemic response and the public health benefits of an effective vaccine.

## Data Availability

The research data are distributed by the USC Dornsife Center for Economic and Social Research.

https://uasdata.usc.edu/index.php

## Author Contributions

Dr Daly had full access to the study data and takes responsibility for the integrity of the data and accuracy of the data analysis.

### Concept and design

All authors.

### Acquisition, analysis, or interpretation of data

All authors.

### Drafting of the manuscript

All authors.

### Critical revision of the manuscript for important intellectual content

All authors.

### Statistical analysis

Daly.

## Acknowledgements

The project described in this paper relies on data from survey(s) administered by the Understanding America Study, which is maintained by the Center for Economic and Social Research (CESR) at the University of Southern California. The content of this paper is solely the responsibility of the authors and does not necessarily represent the official views of USC or UAS. The collection of the UAS COVID-19 tracking data is supported in part by the Bill & Melinda Gates Foundation and by grant U01AG054580 from the National Institute on Aging. However, these organizations bear no responsibility for the analysis or interpretation of the data.

---

**Table S1.** Results of unadjusted multinomial logistic regression analyses examining the relative risk of being undecided or unwilling to vaccinate against COVID-19 in October compared to April, 2020.

| Variable | Undecided on vaccination |  | Unwilling to vaccinate |  |
| --- | --- | --- | --- | --- |
|  | RRR<sup>a</sup> | (95% CI) | RRR<sup>a</sup> | (95% CI) |
| Overall sample | 1.82*** | (1.62, 2.05) | 2.29*** | (2.11, 2.48) |
| Age group |  |  |  |  |
| 18-34 | 1.75*** | (1.34, 2.27) | 2.37*** | (1.98, 2.84) |
| 35-49 | 1.84*** | (1.49, 2.27) | 2.26*** | (1.95, 2.62) |
| 50-64 | 1.92*** | (1.56, 2.35) | 2.54*** | (2.18, 2.95) |
| 65+ |  |  |  |  |
| Sex |  |  |  |  |
| Male | 1.80*** | (1.48, 2.18) | 2.12*** | (1.87, 2.41) |
| Female | 1.90*** | (1.64, 2.20) | 2.48*** | (2.23, 2.76) |
| Race/ethnicity |  |  |  |  |
| White | 1.97*** | (1.72, 2.25) | 2.32*** | (2.12, 2.54) |
| Hispanic | 1.89*** | (1.32, 2.68) | 2.52*** | (2.00, 3.17) |
| Black | 1.41* | (1.03, 1.92) | 2.08*** | (1.62, 2.67) |
| Other race/<br>ethnicity | 3.49*** | (1.94, 6.29) | 2.70*** | (1.79, 4.06) |
| Education |  |  |  |  |
| Degree | 1.71*** | (1.32, 2.21) | 2.09*** | (1.82, 2.41) |
| No degree | 2.01*** | (1.75, 2.31) | 2.52*** | (2.28, 2.79) |
| Income level<sup>b</sup> |  |  |  |  |
| Low income | 2.01*** | (1.70, 2.37) | 2.53*** | (2.20, 2.92) |
| Middle income | 1.83*** | (1.48, 2.25) | 2.17*** | (1.92, 2.46) |
| High income | 1.70*** | (1.18, 2.42) | 2.31*** | (1.93, 2.77) |
| Chronic condition<sup>c</sup> |  |  |  |  |
| No | 1.77*** | (1.53, 2.05) | 2.23*** | (2.02, 2.46) |
| Yes | 1.93*** | (1.59, 2.34) | 2.48*** | (2.15, 2.86) |

<sup>a</sup> RRR = Relative risk ratio. For all analyses “willing to vaccinate” was the outcome reference group.
<sup>b</sup> Households earning less than \$40,000 a year classified as low income, those earning \$40,000 - \$100,000 middle income, and those above this threshold as high-income.
<sup>c</sup> Diagnosed with any of the following: diabetes, cancer, heart disease, kidney disease, asthma, chronic lung disease, an autoimmune disease.
\* $P < .05$ . \*\* $P < .01$ . \*\*\* $P < .001$

